# Evaluation of ultrasonic renal volume in relation to body size in patients with hypertension: Comparative cross-sectional study

**DOI:** 10.1101/2020.08.27.20183079

**Authors:** Elias Kedir, Melkamu Berhane, Tilahun Alemayehu Nigatu, Almaz Ayalew, Mesfin Zewdu

## Abstract

**Introduction:** Estimation of renal size is useful parameter in the diagnosis of abnormal structural change on the kidneys due to the adverse effects of chronic diseases like hypertension. This study evaluated renal volume by ultrasound in relation to body size parameters, notably BMI and body surface area in hypertensive and non-hypertensive individuals.

**Methods:** A hospital-based comparative cross-sectional study was conducted from February to September 2018 at the Radiology department of the Jimma University Medical Center (JUMC). The study included consecutively selected samples of ambulatory hypertensive patients and non-hypertensive controls recruited consecutively on voluntary basis. After providing verbal informed consent, each subject underwent abdominal ultrasound examination; length, width and thickness of both kidneys were measured and used for estimation of renal volume. The statistical evaluation included independent samples t-tests for mean differences with regard to ultrasonic renal measurements between hypertensive and non-hypertensive groups,

**Results:** A total of 145 adults aged 16 - 80 years (mean ±SD=44 ±17) participated in the study. In the hypertensive group, renal volume ranged 36.1 - 201.6 (mean=97.7) cm^3^ for the right kidney and 39.6 - 189.5 (mean=104.4) cm^3^ for the left kidney, whilst it was 61.8 - 159.5 (mean=101.1) cm^3^ for the right and 35.8 - 253.7 (mean=111.8) cm^3^ for the left kidney among the control group. Both kidneys were slightly smaller in the hypertensive group as compared to the controls. Right renal volume to BSA ratio ranged from 23.5 - 100.6 (mean=58.2) cm^3^/m^2^ in hypertensive group, while it was between 37.0 and 96.1 (mean=62.6) cm^3^/m^2^ among the control group (p=0.076). Left renal volume to BSA of the patients which ranged from 24.1 - 97.1 (mean=62.2) cm^3^/m^2^ was significantly (p=0.012) lower than that of the non-hypertensive group, which was between 23.6 and 132.5 (mean=69.3) cm^3^/m^2^.

**Conclusion:** The results of this study have shown slightly smaller bilateral renal volume among hypertensive patients as compared the controls. We recommend large scale research in other parts of Ethiopia so that nationally representative data can be obtained.

## Introduction

Hypertension, defined as persistently elevated blood pressure (BP), is a multifactorial non-communicable disorder that substantially contributes to the global burden of diseases. Hypertension is a well-known modifiable risk factor for several illnesses including renal failure [1], cardiovascular diseases [2] and premature death worldwide [3]. In the recent past, the prevalence and absolute burden of hypertension has raised globally, especially in low- and middle-income countries (LMICs), including Ethiopia [4, 5].

The kidneys are among the organs commonly affected by hypertension, hence critical targets of hypertension-induced organ damage [6]. Understanding the early stages of the interaction between blood weight and renal work is basically vital for prevention of hypertension and associated renal malady. A distant better understanding of the impacts of basic hypertension on renal work may offer assistance for early location of the illness, follow-up, and to prompt treatment on evidence base [7].

Estimation of renal size could be a crucial step in the evaluation and treatment of long standing illnesses like hypertension [8]. Renal size estimation most commonly incorporates renal length, volume and cortical thickness [9]. For ordinary hone, renal length estimation is more solid because of its simple reproducibility, but most precise is the renal volume estimation [10]. Additionally, the foremost exact estimation of renal state is the whole renal volume related with height, weight, and added up to body surface area (BSA) [11]. In clinical practice, BSA approximates total surface area of the body and is used to calculate drug dosages and as an indicator of the health status of individuals [12].

Demonstrative imaging modalities and strategies such as ordinary radiography (CR), computed tomography (CT), attractive reverberation imaging (MRI), atomic medication (NM), and ultrasonography among others have been utilized for renal assessment, particularly in terms of estimate and work, but no single strategy is generally acknowledged for renal estimate appraisal [13-15]. Even though different imaging modalities are available to be used for renal volume assessment, ultrasonography (US) has replaced standard radiography and has become the standard imaging modality in the investigation of renal diseases due to its noninvasive nature and easy availability [7]. Additionally, it offers excellent anatomical details, doesn’t require special patient preparation and does not expose patients to radiation or contrast agents.

Different studies have shown that anthropometric estimations like height, weight, and body mass index (BMI) relate exceptionally well with renal length and volume [16, 17]. Higher BMI is associated with increased risk of several non-communicable diseases like diabetes mellitus and hypertension, which if not treated timely and properly can lead to end-stage renal disease (ESRD) [18].

Kidney size measurements have traditionally been used as predictors of chronic kidney diseases; however, these predictions are often based on an incomplete knowledge of accuracy and evolving evidence of effectiveness. Kidney length may not be an absolute predictor of overall kidney size, perhaps due in part to the fact that it measures only a single renal dimension, which is subject to inconsistency pertaining considerably to the varied shape of the kidneys within or between individuals. Renal volume (RV) rather, has been emphasized by several authors as a true predictor of kidney size in states of good health and disease [19, 20].

There is no study done in Ethiopia on renal size measurements as determinant parameters either in healthy people or in those with conditions such as hypertension, diabetes mellitus, and renal disease. Therefore, this study was done with the objectives of evaluating renal volume in patients with hypertension and correlate it with anthropometric parameters as compared to non-hypertensive controls.

## Materials and methods

### Study area, design and subjects

A hospital-based comparative cross-sectional study was conducted from February to September 2018 at the Radiology Department of the Jimma University Medical Center (JUMC). The study participants were consecutively selected samples of hypertensive patients and non-hypertensive controls. The cases were patients with hypertension who have been on follow up at JUMC chronic illnesses follow up clinic, whereas the controls were apparently healthy hospital visitors who have no known history of renal diseases, hypertension or diabetes. After obtaining informed verbal consent, each study subject underwent abdominal ultrasound of both kidneys.

### Participants’ inclusion and exclusion criteria

Inclusion criteria for the hypertensive group were being 16 years or older and on regular follow-up for ≥1 year for established hypertension with no history of renal disease whereas for the controls, it was age ≥16 years and no any history of hypertension, diabetes mellitus or renal diseases. Additionally, presence of bilateral, grossly symmetric kidneys on US verified fulfilment of inclusion criteria of the subjects [21]. People with chronic renal disease, pregnant women and women who have given birth in the last 12 months were excluded. Further, subjects with ultrasonic evidences of abnormal kidneys such as horseshoe or ectopic kidney and/or those with renal cysts were also excluded from final analysis.

### Ultrasonic examination and somatic measurements

Participants in both study groups underwent abdominal US examination with the same US machine (General Electric Health care LOGIQ P6, B-Model) using the 4 MHz curvilinear probe. Each subject had scanning of both kidneys in supine and decubitus positions in the longitudinal and transverse planes for renal length, width and antero-posterior (AP) diameter (thickness) in centimeters. The liver and spleen were used as acoustic windows for the right and left kidneys respectively [22]. No prior preparations of study subjects were required before examination. Renal length (RL) was taken on a coronal scan as the longest distance between the superior and inferior poles of the kidney using an electronic caliper. The AP diameter (thickness) was measured on a sagittal scan as the maximum distance between the anterior and posterior walls at the mid-third of the organ. The renal width (W) was measured on a transverse scan as the longest distance between the medial and lateral borders away from the hilum of the kidney. These three measurements were later used to estimate overall renal volume (RV) of the ipsilateral kidney.

Participants were first interviewed for completed age, sex and duration of hypertension in years since diagnosis and history of kidney problems. The height (H) in meters and weight (W) in kilograms of the subjects were measured while standing erect against a ZT World Health Organization (WHO) weighing scale, and used for BMI and BSA calculations.

### Outcome measures

The main outcome variable in this study was bilateral renal volume (RV), which was derived from the three absolute ultrasonic renal dimensions measured. On each side, renal volume was computed electronically on statistical software using an ellipsoid formula RV= RL × W × AP × 0.523 as originally described by Hricak and Lieto (1983) [23]. Other variables include BMI and BSA, both derived from body weight (W) and height (H). BMI was estimated as a ratio of weight in kg to height in meter squared. Body surface area was computed using the Mosteller formula that takes the square root of the height (m) multiplied by the weight (kg) divided by 36 [12, 24]. To account for general body physique variation among individuals with respect to renal size, renal volume to surface area ratio (RV/BSA) was also computed arithmetically as additional study variable.

### Data processing and analysis

Collected data were checked for completeness and error, then coded and entered into Statistical Package for Social Sciences (SPSS) for windows version 23 [25]. Preliminary inspection of the numerical data included minimum, maximum, mean, standard deviation (SD), median and interquartile range (IQR). The statistical evaluation included independent samples t-tests for mean differences with regard to age, somatic and ultrasonic renal measurements between hypertensive and non-hypertensive groups, as well as between male and female subjects. The renal sizes on the two sides of the body were also compared with pair-sample t-tests. Bivariate correlations of the renal volume with age, body weight, height, BMI and BSA were assessed using Pearson’s Product correlation coefficient (*r*), separately for the two study groups. All statistical tests were two-tailed and considered significant at *p*<0.05.

### Ethical approval

Ethical approval was obtained from the Ethical Review Board of Jimma University, Institute of Health. Formal permission was also sought from the hospital administration and radiology department. Before enrolment, participants were informed about the study purpose and requested for their interest to participate in the study. Those who agreed and provided voluntary verbal consent were included in the study.

## Results

### Main characteristics of the study participants

A total of 145 adults (74 males and 71 females) participated in the study; 85 hypertensive outpatients (40 males and 45 female), and 60 (34 male and 26 female) non-hypertensive controls. Self-reported duration of hypertension since diagnosis ranged from 1 to 24 completed years, with a mean duration of 7. The age of the participants ranged from 16 - 80 with a mean (±SD) of 44 (±17) years. The mean BMI and BSA were 22.3 kg/m^2^ (range: 14.4 - 37.3) and 1.65 m^2^ (range: 1.25 - 2.09) respectively (Table 1). With regard to renal size, the RRV ranged from 36.1 to 201.6 cm^3^ (mean=99.1), while LRV ranged from 35.8 to 253.7 cm^3^ (mean=107.4). The RRV/BSA ranged from 24.53 to 100.7 (mean=60.0) cm^3^/m^2^, while LRV/BSA ranged from 23.5 to 132.5 (mean=65.1) cm^3^/m^2^. Both renal volume parameters were significantly different (p<0.01) between the right and left kidneys, the left kidney being larger than the right (Table 1).

**Table 1.** Main characteristics of study participants, Jimma University Medical Center (JUMC), Jimma, Southwest Ethiopia, 2018.

| Variables (Valid N= 145) | Mean | SD | Min. | Q1, 25% | Q2, Median | Q3, 75% | Max. |
| --- | --- | --- | --- | --- | --- | --- | --- |
| Age (year) | 44.4 | 17.3 | 16.0 | 28.0 | 46.0 | 58.0 | 80.0 |
| Body weight (kg) | 59.96 | 12.0 | 37.3 | 50.9 | 68.1 | 76.0 | 96.0 |
| Body height (m) | 1.640* | 0.092 | 1.44 | 1.440 | 1.850 | 1.700 | 1.85 |
| BMI ($\text{kg}/\text{m}^2$) | 22.32* | 4.30 | 14.39 | 21.50 | 22.00 | 24.93 | 37.3 |
| BSA ($\text{m}^2$) | 1.646* | 0.184 | 1.247 | 1.508 | 1.635 | 1.768 | 2.09 |
| Right renal length (cm) | 9.598 | 0.957 | 6.970 | 8.035 | 9.570 | 10.375 | 11.8 |
| Left renal length (cm) | 9.570 | 0.892 | 6.950 | 9.100 | 9.550 | 10.070 | 12.0 |
| Right renal width (cm) | 4.956* | 0.595 | 3.380 | 4.515 | 5.00 | 5.390 | 6.30 |
| Left renal width (cm) | 3.956 | 0.652 | 1.630 | 4.485 | 4.970 | 5.390 | 7.20 |
| Right renal thickness (cm) | 3.908<sup>a</sup> | 0.540 | 2.700 | 3.460 | 3.900 | 4.200 | 5.70 |
| Left renal thickness (cm) | 4.242<sup>a</sup> | 0.578 | 2.820 | 2.830 | 4.200 | 4.620 | 6.18 |
| Right renal volume ($\text{cm}^3$) | 9.115<sup>b*</sup> | 28.160 | 36.073 | 80.499 | 96.984 | 118.577 | 201.5 |
| Left renal volume ($\text{cm}^3$) | 107.416<sup>b</sup> | 31.410 | 35.828 | 88.456 | 106.530 | 128.057 | 253.6 |
| RRV/BSA ($\text{cm}^3/\text{m}^2$) | 60.008<sup>c</sup> | 14.678 | 24.525 | 50.116 | 58.908 | 68.359 | 100.6 |
| LRV/BSA ($\text{cm}^3/\text{m}^2$) | 65.130<sup>c</sup> | 16.967 | 23.522 | 53.755 | 65.812 | 74.307 | 132.4 |

<sup>a,b,c</sup>, the mean values in the row are statistically significant for the right and left kidneys; Min., minimum; Max., maximum; BMI, body mass index; BSA, body surface area; Q1, first quartile; Q2, second quartile, Q3, third quartile; SD, standard deviation; LRV, left renal volume; RRV, right renal volume; \*the mean scores are significantly different between male and female.

### Comparison of hypertensive and non-hypertensive groups

Table 2 shows comparison of the two study groups disaggregated by sex with regard to their renal size and other variables. The mean age of the non-hypertensive group was 33 (range: 16-80) years, while that of the hypertensives was 53 (range: 20-78) years with no age difference between male and female subjects in both groups. Overall, the mean BMI was significantly higher in hypertensive group (mean= 23.4 kg/m^2^) than non-hypertensive group (20.9 kg/m^2^) in both sexes (Table 2).Table 2. Comparison of ultrasonic renal volume and somatic variables between hypertensive patients and non-hypertensive controls stratified by sex, Southwest Ethiopia 2018.

**Table 2.** Comparison of ultrasonic renal volume and somatic variables between hypertensive patients and non-hypertensive controls 183 stratified by sex, Southwest Ethiopia 2018.

| Variable | Non-hypertensive controls ( N=60) |  |  |  |  |  | Hypertensive patients (N=85) |  |  |  |  |  | p-value |
| --- | --- | --- | --- | --- | --- | --- | --- | --- | --- | --- | --- | --- | --- |
|  | Sexes | n | Min. | Max. | Mean | SD | n | Min. | Max. | Mean | SD | □-<br>statistic |  |
| Age (year) | F | 26 | 16.0 | 80.0 | 33.19 | 16.46 | 45 | 22.0 | 70.0 | 50.1 | 12.52 | -4.878** | 0.000 |
|  | M | 34 | 18.0 | 63.0 | 32.23 | 13.25 | 40 | 20.0 | 78.0 | 55.75 | 15.06 | -7.070** | 0.000 |
|  | F + M | 60 | 16.0 | 80.0 | 32.65 | 14.60 | 85 | 20.0 | 78.0 | 52.76 | 13.98 | -8.377** | 0.000 |
| Body weight (kg) | F | 26 | 37.3 | 80.0 | 56.71 | 11.37 | 45 | 41.0 | 96.0 | 61.70 | 14.47 | -1.507 | 0.136 |
|  | M | 34 | 43.8 | 77.3 | 57.20 | 7.66 | 40 | 45.0 | 86.0 | 62.46 | 11.86 | -2.297 | 0.025 |
|  | F + M | 60 | 37.3 | 80.0 | 56.99 | 9.36 | 85 | 41.0 | 96.0 | 62.06 | 13.24 | -2.701* | 0.008 |
| Body height (m) | F | 26 | 1.44 | 1.76 | 1.58<sup>a</sup> | 0.07 | 45 | 1.47 | 1.80 | 1.58<sup>c</sup> | 0.06 | -0.019 | 0.985 |
|  | M | 34 | 1.52 | 1.85 | 1.72<sup>a</sup> | 0.07 | 40 | 1.50 | 1.82 | 1.69<sup>c</sup> | 0.07 | 1.746 | 0.085 |
|  | F + M | 60 | 1.44 | 1.85 | 1.66 | 0.10 | 85 | 1.47 | 1.82 | 1.63 | 0.08 | 1.709 | 0.090 |
| BMI (kg/m<sup>2</sup>) | F | 26 | 14.39 | 29.38 | 22.70<sup>b</sup> | 3.72 | 45 | 19.0 | 37.32 | 24.65<sup>d</sup> | 4.93 | -1.750 | 0.084 |
|  | M | 34 | 15.47 | 25.46 | 19.46<sup>b</sup> | 2.53 | 40 | 16.56 | 32.39 | 21.88<sup>d</sup> | 3.59 | -3.389* | 0.001 |
|  | F + M | 60 | 14.39 | 29.38 | 20.86 | 3.47 | 85 | 16.56 | 37.32 | 23.35 | 4.54 | -3.562** | 0.000 |
| BSA (m<sup>2</sup>) | F | 26 | 1.23 | 1.92 | 1.57 | 0.18 | 45 | 1.32 | 2.09 | 1.64 | 0.21 | -1.316 | 0.192 |
|  | M | 34 | 1.41 | 1.94 | 1.65 | 0.12 | 40 | 1.42 | 2.01 | 1.71 | 0.18 | -1.597 | 0.115 |
|  | F + M | 60 | 1.25 | 1.94 | 1.61 | 0.15 | 85 | 1.32 | 2.09 | 1.67 | 0.20 | -1.829 | 0.069 |
| RRV (cm<sup>3</sup>) | F | 26 | 61.75 | 159.5 | 96.41 | 24.53 | 45 | 45.81 | 163.9 | 92.63 | 23.52 | 0.643 | 0.522 |
|  | M | 34 | 63.72 | 152.7 | 104.68 | 22.65 | 40 | 36.07 | 201.6 | 103.44 | 36.68 | 0.176 | 0.861 |
|  | F + M | 60 | 61.75 | 159.5 | 101.10 | 24.20 | 85 | 36.07 | 201.6 | 97.72 | 30.72 | 0.711 | 0.478 |
| LRV (cm<sup>3</sup>) | F | 26 | 35.83 | 253.7 | 105.21 | 40.67 | 45 | 53.71 | 162.7 | 102.9 | 25.64 | 0.294 | 0.770 |
|  | M | 34 | 77.06 | 169.1 | 116.76 | 22.40 | 40 | 39.56 | 189.5 | 105.98 | 36.20 | 1.556 | 0.122 |
|  | F + M | 60 | 35.83 | 253.7 | 111.76 | 31.86 | 85 | 39.56 | 189.5 | 104.35 | 30.91 | 1.405 | 0.162 |
| RRV/BSA (cm<sup>3</sup>/m<sup>2</sup>) | F | 26 | 41.9 | 96.1 | 61.48 | 14.33 | 45 | 33.62 | 99.1 | 56.58 | 12.31 | 1.520 | 0.133 |
|  | M | 34 | 37.0 | 93.4 | 63.43 | 13.01 | 40 | 23.53 | 100.6 | 60.01 | 18.01 | 0.924 | 0.358 |
|  | F + M | 60 | 37.0 | 96.1 | 62.58 | 13.51 | 85 | 23.53 | 100.6 | 58.19 | 15.26 | 1.789 | 0.076 |
| LRV/BSA<br>(cm<sup>3</sup>/m<sup>2</sup>) | F | 26 | 23.52 | 132.5 | 67.10 | 22.85 | 45 | 38.6 | 97.4 | 63.04 | 14.64 | 0.905 | 0.369 |
|  | M | 34 | 41.5 | 104.1 | 71.01 | 13.10 | 40 | 24.05 | 94.7 | 61.23 | 16.99 | 2.734* | 0.008 |
|  | F + M | 60 | 23.62 | 132.5 | 69.30 | 17.92 | 85 | 24.05 | 97.4 | 62.19 | 15.71 | 2.532* | 0.012 |

F, female; M, male; BMI, body mass index; BSA, body surface area; Max, maximum; Min, minimum; SD, standard deviation; <sup>a,b,c,d,e</sup>, the values indicated are statistically significant for the right and left kidneys; LRV, left renal volume; RRV, right renal kidney volume \*significant at p<0.01; \*\*significant at p<0.001 between men and women

In the hypertensive group, renal volume of both sexes ranged from 36.1 to 201.6 (mean=97.7) cm^3^ for the right kidney and 39.6 to 189.5 (mean=104.4) cm^3^ for the left kidney. In this group, mean volumes of the right and left kidneys in males were 103.4 (±36.7) and 106.0 (±36.2) cm^3^ respectively, while it was 92.6 (±23.5) and 102.9 (±25.6) cm^3^ respectively for females (Table 2). In the non-hypertensive group, the renal volume ranged from 61.8 to 159.5 (mean=101.1) cm^3^ for the right and from 35.8 to 253.7 (mean=111.8) cm^3^ for the left kidney, indicating slightly larger kidneys on both sides in this group as compared to the hypertensive group.

When renal volume on each side is seen in terms of body surface area, RRV/BSA ranged from 23.5 to 100.6 (mean=58.2) cm^3^/m^2^ in the hypertensive group, while it was between 37.0 and 96.1 (mean=62.6) cm^3^/m^2^ among the non-hypertensive group (p=0.076). In contrary, LRV/BSA of the hypertensive group ranging from 24.1 to 97.1 (mean=62.2) cm^3^/m^2^ was significantly (p=0.012) lower than that of the non-hypertensive group, which was 23.6-132.5 (mean=69.3) cm^3^/m^2^ (Table 2).

### Factors associated with renal volume

Relationship of the RRV and LRV with age, weight, height, BMI and BSA was shown in Table 3. As shown, neither the right nor the left renal volume has significant correlation with age in either group or sex. The largest mean renal volumes for right and left kidney were recorded in the same age group (40-49 years) in the male and female hypertensive subjects, in the control group however largest renal volumes were calculated for those in the fourth decades (30—39 yrs). As depicted in Table 3, on both sides. BMI and BSA strongly correlated with renal volume, particularly among the hypertensive patients.

**Table 3.** Pearson correlations between renal volume and somatic parameters in male and female hypertensive patients and controls^a^.

| Variable | Sex | Hypertensive patients |  | Non-hypertensive controls |  |
| --- | --- | --- | --- | --- | --- |
|  |  | Right RV | Left RV | Right RV | Left RV |
| Age | Female | 0.132 | 0.083 | -0.127 | -0.011 |
|  | Male | -0.158 | -0.179 | -0.016 | -0.194 |
|  | Both sexes | -0.011 | -0.058 | -0.075 | 0.057 |
| Body weight | Female | 0.538** | 0.459** | 0.381 | 0.328 |
|  | Male | 0.583** | 0.698** | 0.364* | 0.223 |
|  | Both sexes | 0.531** | 0.560** | 0.366** | 0.293* |
| Body height | Female | 0.372* | 0.231 | 0.267 | 0.145 |
|  | Male | 0.547** | 0.646** | 0.391* | 0.128 |
|  | Both sexes | 0.472** | 0.395** | 0.353* | 0.222 |
| BMI | Female | 0.463** | 0.436** | 0.288 | 0.295 |
|  | Male | 0.367* | 0.459** | 0.150 | 0.187 |
|  | Both sexes | 0.308* | 0.383* | 0.118 | 0.140 |
| BSA | Female | 0.548** | 0.456** | 0.392* | 0.307 |
|  | Male | 0.620** | 0.743** | 0.423* | 0.234 |
|  | Both sexes | 0.576** | 0.587** | 0.425** | 0.313* |

<sup>a</sup>values are Pearson's correlation coefficients; BMI, body mass index; BSA, body surface area; \*\*correlation is significant at the $p < 0.01$ level (2-tailed); \*correlation is significant at the $p < 0.05$ level (2-tailed).

In hypertensive patients, renal volume was correlated significantly (p<0.05) with BMI (r=0.308 and 0.383 for right and left kidneys, respectively). Further, significant positive correlation was seen between renal volume and BSA in the hypertensive group r =0.576 and 0.587 (p<0.01) for the right and left kidneys respectively. When stratified by sex, these correlations were still strong and significant (Table 3). Among non-hypertensive controls, in contrast, only BSA showed significant correlation with renal volume on both sides in both sexes (Table 3).

## Discussion

In the recent past, the prevalence and absolute burden of the well-known modifiable risk factor of renal failure, hypertension [1], has raised globally especially in LMICs, including Ethiopia [2–5]. Currently, ultrasonic renal size reports from Ethiopia are scarce [26].

Although ultrasonic length, width and thickness of both kidneys were measured for each study participant, computed renal volume was used as a proxy measure of overall kidney size in this study. Preference of renal volume to individual renal dimension is dual imperative. Firstly, the usual bean-shape of each kidney is subject to considerably varied shape and orientation [27]. This could potentially lead to erroneous recording of the three dimensions (length, width and thickness) such as exchange of one dimension for the other. Relying on only one dimension, for instance renal length, for estimation of overall renal size could therefore arrive at significant risk of wrong conclusion and recommendation about kidney status. Secondly, renal volume is three-dimensional modality estimated from records of organ size from three scanning planes. As is composite outcome measure incorporating all the three dimensions, renal volume is believed to be the more realistic predictor of actual size. As a result, ultrasonic renal volume is increasingly entering the repertoire of kidney size evaluation in clinical practice [19, 20].

The result shows that, the renal volume of the hypertensive group for the left kidney was slightly smaller than the size calculated for non-hypertensive controls. The renal volume obtained in the current population is comparable with results reported from Sudan among similar study groups [28], but smaller than that reported from Nigeria [19], possibly due to the differences in the study population.

In clinical practice, bilaterally shrunken kidney as a result of chronic disease supports the diagnosis of chronic kidney disease [21]. In our study, we observed slightly smaller bilateral renal volume among hypertensive patients as compared to non-hypertensives. However, the difference was small and not statistically significant. This finding is in agreement with a report from Turkey [7], which also reported reduced renal volume in hypertensive patients when compared with non-hypertensive individuals.

The small sample size was one of the limitations of our study. Further, while attempting to provide insights on the impact of hypertension on the kidneys in this study, the approach focused only on anatomical aspects i.e. ultrasonic renal size, regardless of pathophysiologic considerations.

## Conclusions

The renal volume of both kidneys was found smaller than that reported from Africa and the rest of the world in both study groups. Moreover, the renal size was slightly smaller among hypertensive patients as compared their control counter parts. We recommend large scale research including other regions of Ethiopia so that we will have fully standardized data on the subject.

## Data Availability

All relevant data are within the manuscript and its Supporting Information files.

## Acknowledgments

We thank all the staff of JUMC for their assistance and cooperation during the data collection process. We also thank Jimma University Institute of Health for financially supporting this study. Finally, we are grateful to the study participants for their cooperation.

## References

1. Sim JJ, Shi J, Kovesdy CP, Kalantar-Zadeh K, Jacobsen SJ. Impact of achieved blood pressures on mortality risk and end-stage renal disease among a large, diverse hypertension population. J. Am College Cardio 2014; 6 4 (6). ISSN 0735-1097 /$36.00. Available at http://dx.doi.org/10.1016/j.jacc.2014.04.065.

2. Kotchen TA. Hypertensive vascular disease, In Kasper, DL, Hauser SL, Jameson JL, Fauci AL, Longo DL, Loscalzo J, editors. Harrison’s Principles of Internal Medicine, 19^th^ edition, 2015. McGraw-Hill Education Publishing Inc, New York, PP. 1611-1626, ISBN: 978-0-07-180216-1.

3. Mills KT, Stefanescu A, He J. The global epidemiology of hypertension. Nat Rev Nephrol. 2020; 16: 223–237.

4. Abebe S, Yallew WW. Prevalence of hypertension among adult outpatient clients in hospitals and its associated factors in Addis Ababa, Ethiopia: a hospital based cross-sectional study. BMC Res Notes, 2019; 12:87. https://doi.org/10.1186/s13104-019-4127-1.

5. Roba HS, Beyene AS, Mengesha MM, Ayele BH. Prevalence of hypertension and associated factors in Dire Dawa City, Eastern Ethiopia: A community-based cross-sectional study. Hindawi Intern J Hypertension. 2019: Article ID 9878437.

6. Singh GR, Wendy EH. Kidney volume, blood pressure, and albuminuria; findings in an Australia aboriginal community. Am J Kidney Dis. 2004; 43: 254–259.

7. Zumrutdal AO, Turan C, Cetin F, Adanali S. Relationship between renal size and hypertension in a patient with CRF. Nephron. 2002; 90: 1457–1462.

8. Muthusamy P, Ananthakrishnan R, Santosh P. Need for a nomogram of renal sizes in the Indian population. Findings from a single-center sonographic study. Indian J Med Res. 2014; 139: 686–639.

9. Saeed Z, Mirza W, Sayani R, Sheikh A, Yazdani I, Hussain SA, Sonographic measurement of renal dimensions in adults and its correlates, Int. J. Collab. Res. Intern. Med. Public Health 2012; 4 (9): 1626–1641.

10. Abdullah MB, Garelnabi MB, Ayad CE, Abdalla EA. Establishment of reference values for renal length and volume for normal adult Sudanese using MRI disc summation method. Global J. Medi. Res. 2014; 14: 29–37.

11. Maravi P, Khan M, Kaushal L, Goyal S. Renal volumes by ultrasound and its correlation with body mass index and body surface area in adult population. Tropical J of Radiology and Imaging 2019; 1(1): 20-26.

12. Mosteller RD. Simplified Calculation of Body Surface Area. N Engl J Med 1987 Oct 22; 317 (17): 1098 (letter). *[Cross-referenced by Schnur, M.B*., 2017].

13. Eze CU, Marcie TT. Ultrasonographic kidney sizes among children in Benin, Nigeria: correlation with age and BMI. Radiol. Technol 2013; 84: 341–347.

14. Jeffri A, Abdulla A. Ultrasonographic measurement of kidney Dimensions. Acta Medica Philipina. 2010; 44: 35–38.

15. Maaji SM, Daniel O, Adamu B. Sonographic measurement of renal dimensions of adults in northwestern Nigeria: a preliminary report. Sub-Saharan Afr. J. Med. 2015; 2: 123–7.

16. Kang KY, Lee YJ, Park SC, et al. A comparative study of methods of estimating kidney length in kidney transplantation donors. Nephrol Dial Transplant.2007; 22: 2322-2327.

17. Soheilipour F, Jesmi F, Rahimzadeh N et al. Configuring a better estimation of kidney size in obese children and adolescents. Iran J Pedi. 2016; 26: e4700.

18. Ren Q, Su C, Wang H, et al. Change in BMI and its impact on the incidence of hypertension in 18-65-year-old Chinese adults. PloS One. 2016; 13: 257.

19. Adedeji A, Egberongbe, Victor A, et al. Evaluation of renal volume by ultrasonography in patients with essential hypertension in Ile-Ife, southwestern Nigeria. Libya. J. Medi. 2010; 5: 4848–4869.

20. Sanusi AA, Arogundade FA, Famurewa OC, Akintomide AO, Soyinka FO, Ojo OE et al., Relationship of ultrasonographically determined kidney volume with measured GFR, calculated creatinine clearance and other parameters in chronic kidney disease, Nephrol. Dial. Transplant. 2009; 24: 1690–1694. https://doi.org/10.1093/ndt/gfp055.

21. Bargman JM, Skorecki K, Chronic kidney disease, in: D.L. editions of Kasper SL, Hauser JL, Jameson AL, Fauci DL, Loscalzo J (Eds.): Harrison’s Principles of Internal Medicine, nineteenth ed., McGraw-Hill Education, NewYork, 2015, p. 1811—1921.

22. Gamal AbS, Esam M, Ali MA. Ultrasonographic estimation of age-dependent changes in length of spleen and hepatic lobes and diameters of portal vein and common bile duct in children. J Am Sci. 2013; 9(11):31-39. http://www.jofamericanscience.org.

23. Hricak H, Lieto RP. Sonographic determination of renal volume. Radiology. 1983; 148: 311–2.

24. Schnur MB, 2017. Body Mass Index and Body Surface Area: What’s the Difference? https://www.nursingcenter.com/ncblog/august-2017/body-mass-index-and-body-surface-area-what-s-the-d (last visited on May 19, 2020).

25. IBM-Corp. Statistical Package for Social Sciences (SPSS) for windows version 23, IBM-SPSS, 2015.

26. Diliab D, Mesfin Z, Almaz A, Tilahun, AN. Ultrasonic renal size and its correlates among diabetic outpatients at Jimma University Medical Center, Southwest Ethiopia. Translational Research in Anatomy. 2020; 20, 100071. https://doi.org/10.1016/j.tria.2020.10007 doi:10.4172/2161-0940.1000246.

27. Abd Elgyoum AM, Osman H, Elzaki A, Abd Elrahim E. Ultrasonography patterns for diabetic nephropathy according to the body shape. Sch. J. App. Med. Sci. 2014; 2(5C):1649-1652.

28. Mohamed EMG, Ahmed A, Hossam G, Adil A, Mansour MH. Characterization of renal function and morphology in hypertensive patients using renal ultrasonography. European Academic Research. 2019; 5(12): 2286-4822.

